## Supplementary figures and images for "Economic Impact of a Precision Nutrition Digital Therapeutic on Employer Health Costs: A Multi-Employer and Multi-Year Claims Analysis"

### Supp Fig S1

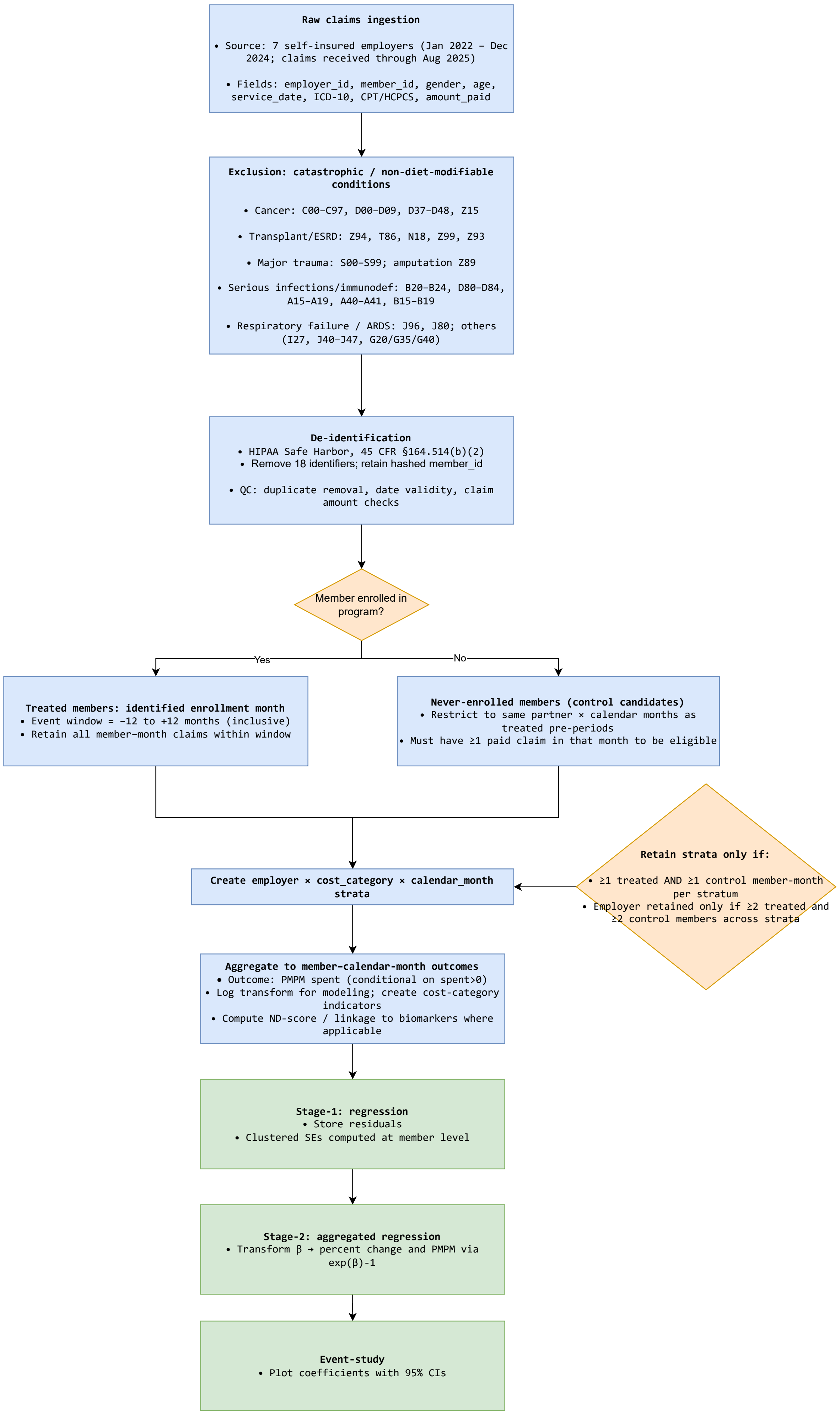
