## Supplementary material for "Economic Impact of a Precision Nutrition Digital Therapeutic on Employer Health Costs: A Multi-Employer and Multi-Year Claims Analysis": Supp Fig S2

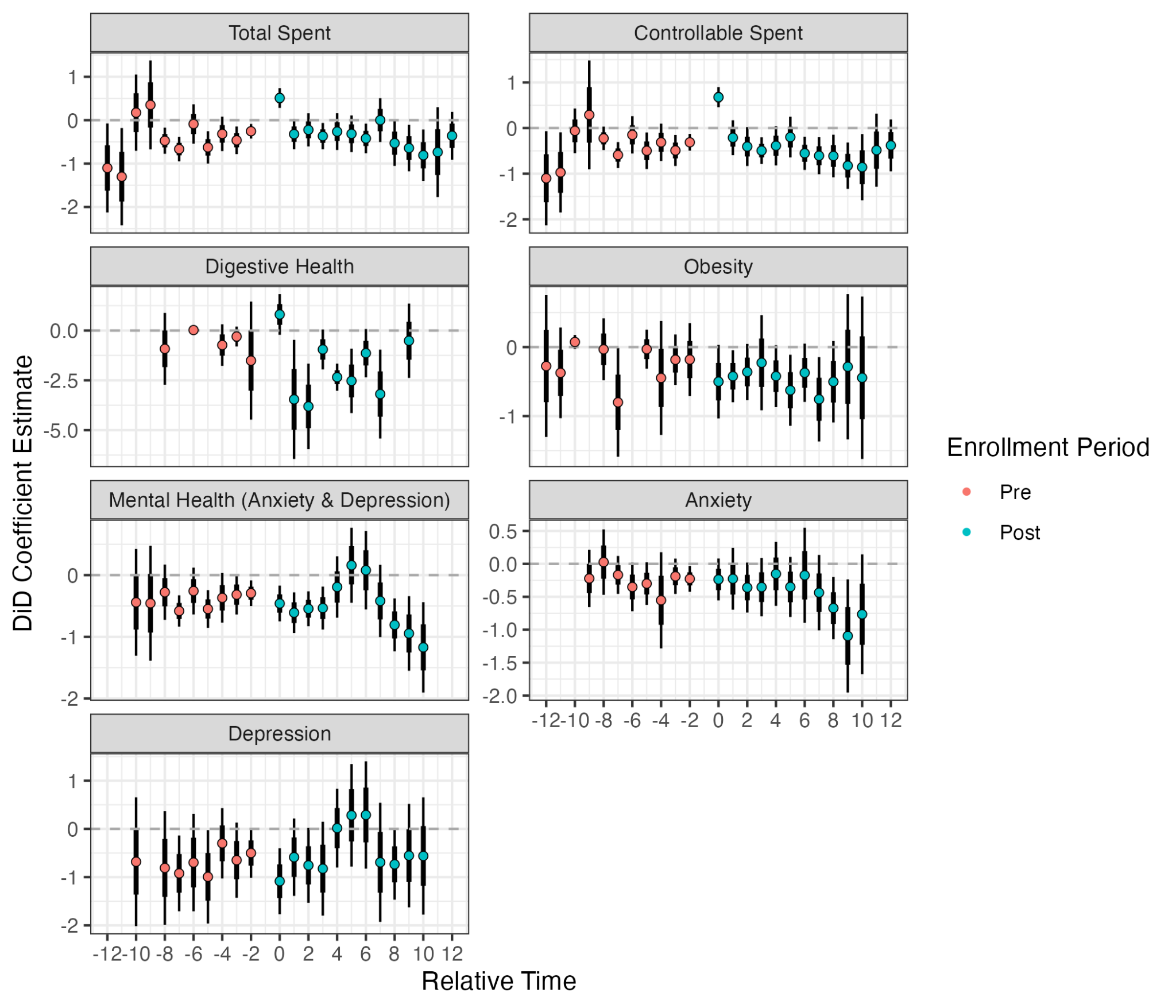

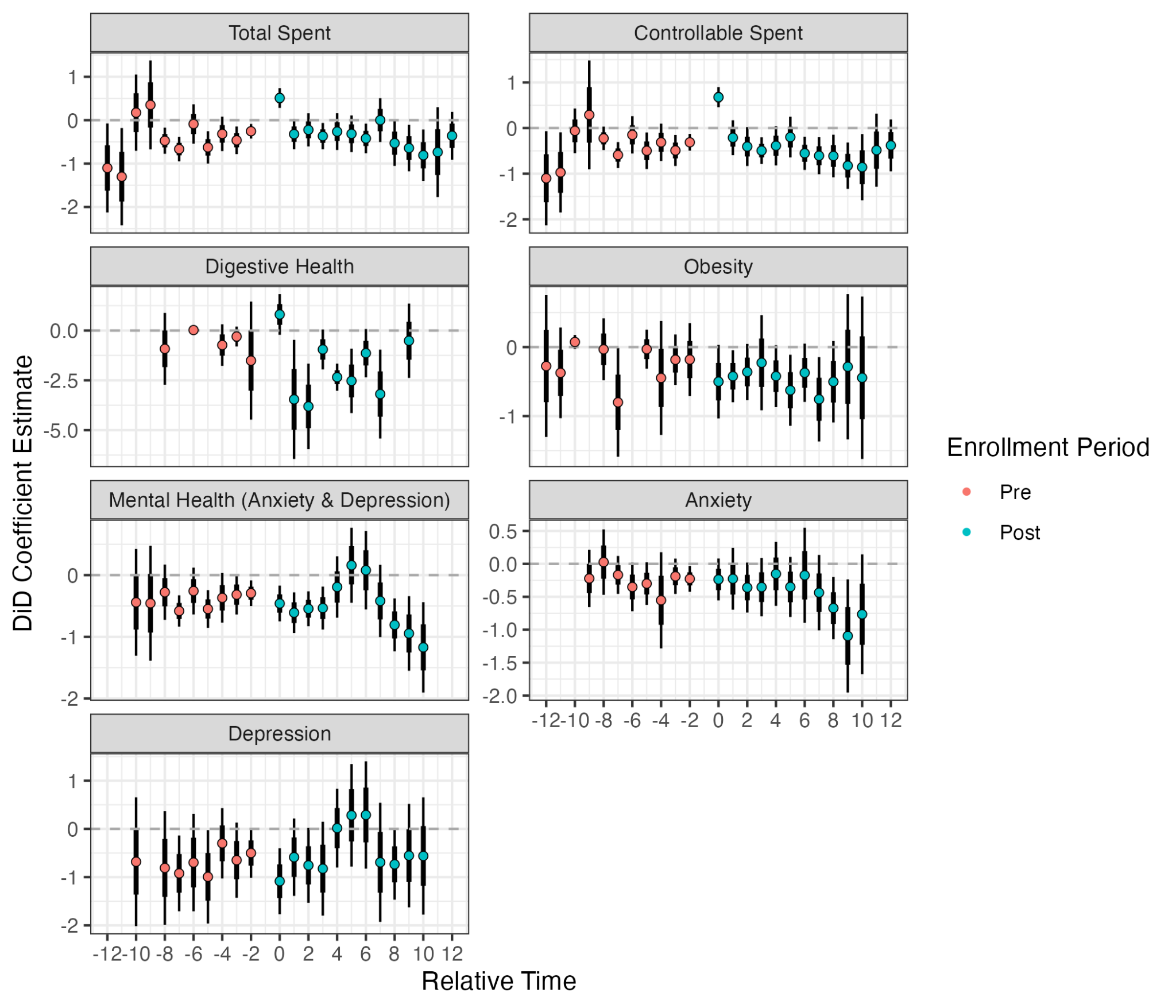

### **Supplementary Figure S1.** Event-study (relative-time) estimates of monthly medical spending before and after program enrollment. Each panel provides the results for each of the disease categories under study. Within each panel plot, the points represent the estimated change in log monthly medical spending (“diet-responsive” cost category) relative to the month immediately preceding enrollment (reference = –1). Estimates were obtained using a two-stage difference-in-differences model, which included member and calendar-month fixed effects, clustered standard errors at the member level, and employer-level geometric weights (see the Methods section for additional details). Shaded bands denote mean estimate ± 1 std error (thick vertical line) and 95 % confidence intervals (thin vertical line). Horizontal dashed line at y = 0 represents the values corresponding to no difference between the two groups. Overall, pre-treatment coefficients fluctuated around zero, indicating broadly parallel spending trends between participants and comparison members. In several condition-specific analyses, a short-term increase in spending was observed at the enrollment month (month 0), likely reflecting partial-month exposure, as claims within the same calendar month as program initiation may include both pre- and post-enrollment periods. Subsequent months showed a progressive decline in medical expenditures, consistent with the main difference-in-differences results presented in Table 1 in the main text.
