## Supplementary material for "Economic Impact of a Precision Nutrition Digital Therapeutic on Employer Health Costs: A Multi-Employer and Multi-Year Claims Analysis": Supp Mat

### Supplementary Materials

##### Supplementary Note S1. Definition of “diet-responsive-cost” ICD-10 codes and exclusion ICD10 and CPT/ HCPCS codes.

###### Definition of diet-responsive Cost ICD10 Codes.

We included in the econometric analyses every claim that carried at least one three-character ICD-10 code judged to be modifiable through Digbi Health’s precision-nutrition and behaviour-change programme. The selection of codes was grounded in three complementary considerations:

1. Material economic burden in working-age, privately-insured populations. Cardiometabolic, digestive and mental-health conditions represented by the chosen ICD-10 blocks account for a disproportionate share of employer health-plan expenditure. For example, the 2024 *Milliman Medical Index* places average family costs at US $32,066, with obesity, diabetes and hypertension identified as the fastest-growing cost drivers (Deana Bell et al., 2024). Limiting the analytic universe to these high-prevalence, high-cost diagnoses maximises the study’s relevance to payer return-on-investment decisions.
2. Strong evidence that outcomes for these conditions are nutrition-responsive.
   - Metabolic disease and obesity (E08-E13, E65-E68). The 2025 ADA *Standards of Care* designate intensive lifestyle and nutrition therapy as first-line treatment for both obesity and type 2 diabetes, citing durable glycaemic and cardiometabolic improvements (Berkowitz et al., 2019; American Diabetes Association Professional Practice Committee et al., 2025b)
   - Hypertension and dyslipidaemia (I10-I16, E78). An AHA 2025 scientific statement concludes that intentional weight-loss interventions “produce clinically meaningful blood-pressure reductions” and are cost-effective adjuncts to pharmacotherapy (Hall et al., 2021)
   - Non-alcoholic fatty-liver disease (K70-K77). A 2024 randomised trial of a mobile app-based lifestyle programme demonstrated significant six-month reductions in hepatic fat and aminotransferases compared with usual care (Kwon et al., 2024)
   - Functional gastrointestinal disorders (K58, R10-R19). Low-FODMAP and Mediterranean dietary patterns achieve symptom relief comparable to pharmacological therapy in irritable-bowel-syndrome RCTs (Halmos et al., 2014; Zalvan et al., 2017; Shah et al., 2021, 2022).
   - Anxiety and Depression, stress-related and somatoform disorders (F40-F48 and F32-F33). Emerging nutritional-psychiatry literature implicates gut-microbiota modulation and micronutrient optimization in ameliorating depressive and anxiety symptoms (Sarris et al., 2015; Jacka, 2017; Chatterton et al., 2018; Firth et al., 2019; Liu et al., 2023).
3. Collectively, this body of evidence substantiates the biological plausibility that Digbi’s intervention can alter clinical trajectories, utilisation patterns and, consequently, payer-paid costs for these codes.
4. Alignment with the external policy landscape for Food-is-Medicine interventions. A 2024 *Health Affairs* state-level simulation estimated that providing medically tailored meals to adults with diet-sensitive conditions (largely contained within our code set) would yield a net national saving of US $32 billion annually and avert 3.5 million hospitalisation (Deng et al., 2025). Restricting the evaluation to diet-responsive ICD-10 blocks therefore mirrors emerging reimbursement frameworks and enhances generalisability to employers contemplating similar nutrition-based benefits.

Finally, narrowing the analytic focus avoids dilution of the treatment signal by diagnoses unlikely to respond to lifestyle modification (e.g. trauma, malignancy). This increases statistical power and internal validity while preserving external relevance to payers’ cost-containment priorities.

###### Definition of ICD10 and CPT/ HCPCS used as claims exclusion criteria

To prevent confounding by extremely high-cost, unrelated, or trajectory-dominating conditions and treatments, we prospectively excluded claims with diagnosis or procedure codes that denote cancer, organ transplantation and complications, renal replacement therapy, major trauma/obstetrics, and other catastrophic or chronic end-stage states. These exclusions (i) avoid inflating baseline or follow-up costs for reasons orthogonal to nutrition/behavioral management, (ii) reduce variance that can mask detectible effects, and (iii) improve comparability across heterogeneous employer populations.

The exclusions were applied to diagnosis codes (ICD-10-CM) and procedure codes (CPT/HCPCS) using regular-expression filters exactly as listed below. Diagnosis-based or procedure-based exclusions remove claims from subsequent analyses.

###### Diagnosis-based exclusions (ICD-10-CM)

Cancer (member-level exclusion)

- Malignant neoplasms: C00-C97 (all subcodes)
- In situ neoplasms: D00-D09 (all subcodes)
- Neoplasms of uncertain or unknown behavior, and selected benign ranges: D37-D48 (all subcodes)
- Genetic susceptibility to malignant neoplasm: Z15 (all subcodes)

Rationale: Active/ongoing cancer care (surgery, chemotherapy, radiation) and oncology work-ups dominate spend/utilization trajectories and are not short-term nutrition-modifiable.

###### Additional catastrophic/complex conditions (claim-level exclusion)

- Organ transplant status and complications: Z94 (all subcodes), T86 (all subcodes)
- Major trauma injuries: all S00-S99 chapters
- Amputation status: Z89 (all subcodes)
- End-stage renal disease/CKD: N18 (all subcodes); Z99 (all subcodes) (device/dependence, incl. dialysis)
- HIV/AIDS: B20-B24
- Primary immunodeficiency: D80-D84
- Pulmonary hypertension: I27 (all subcodes)
- Chronic lower respiratory diseases: J40-J47
- Respiratory failure: J96
- Acute Respiratory Distress Syndrome (ARDS): J80
- Major neurologic disorders: G20 (Parkinson), G35 (MS), G40 (epilepsy) (all subcodes)
- Artificial openings: Z93 (all subcodes)
- Cystic fibrosis: E84 (all subcodes)
- Serious infectious diseases: A15-A19 (tuberculosis); A40-A41 (sepsis); B15-B19 (viral hepatitis); B50-B54 (malaria)

Rationale: These diagnoses signal chronic, high-acuity, or catastrophic pathways likely to overwhelm condition-targeted effects.

###### Procedure-based exclusions (CPT/HCPCS)

Organ and tissue transplantation

- Heart transplant: 33945-33948
- Liver transplant: 47135-47136
- Kidney transplant: 50360, 50365
- Pancreas transplant: 48550, 48552
- Hematopoietic/Bone marrow transplant: 38230, 38240-38241

Cancer treatment

- Chemotherapy administration: 96400-96449, 96450-96459, 96460-96469, 96470-96479, 96480-96489, and 96490-*-96499*
- Radiation therapy (delivery): 77401-77499

Renal replacement therapy

- Dialysis services: 90935, 90937, and 90945-90999. Covers intermittent, prolonged, and related dialysis service families.

Major cardiovascular, trauma, neurologic, and obstetrics procedures (catastrophic cost anchors)

- Major cardiac surgeries: 33500-33899
- Orthopedic/trauma (selected families): 23400-23499 (shoulder), 27500-27599 (femur/tibia), 13100-13699 (complex repairs and related)
- Neurosurgery (broad family): 61000-64999
- Obstetrics & delivery (including common prenatal imaging):
  - Global OB/delivery families: 59000-59999 (broad OB family), specifically 59400-59409, 59410, 59510-59515
  - Prenatal ultrasound: 76801-76828

**Supplementary Note S2. Rationale for condition focus and use of CCSR categories**

This study focused on three condition domains—obesity, digestive health, and mental health—that are here classified as “diet-responsive” due to their strong linkage to diet, lifestyle, and metabolic regulation. The rationale for focusing on these domains is threefold:

1. High economic burden in employer-sponsored plans.
   Obesity, gastrointestinal disorders such as irritable bowel syndrome (IBS) and gastroesophageal reflux disease (GERD), and anxiety-related mental health conditions together account for a disproportionate share of costs in working-age adults. Obesity is associated with billions of dollars in excess annual spending and progression to cardiometabolic disease (Deana Bell et al., 2024; American Diabetes Association Professional Practice Committee et al., 2025a). IBS and GERD are highly prevalent—affecting up to 15-20% of adults—and add approximately $3,000 per member per year in incremental costs (Halmos et al., 2014; Sandhu & Fass, 2018). Anxiety disorders account for rising direct medical costs and indirect productivity losses (Sarris et al., 2015; Jacka, 2017; Opie et al., 2018).
2. Established responsiveness to dietary intervention.
   The evidence base for food-as-medicine is particularly strong for these conditions:
   - Obesity: Lifestyle and dietary therapies are endorsed as first-line treatment (American Diabetes Association Professional Practice Committee et al., 2025a) . Precision nutrition approaches, including microbiome-informed interventions, have been shown to improve weight and metabolic risk (Volpp et al., 2023; Kwon et al., 2024; Ridberg et al., 2025).
   - Digestive health: Dietary modification, particularly low-FODMAP and elimination approaches, is effective in managing IBS and GERD symptoms, improving quality of life, and lowering utilization (Halmos et al., 2014; Sandhu & Fass, 2018).
   - Mental health (Anxiety and Depression): Nutritional psychiatry trials such as SMILES and HELFIMED show dietary interventions reduce depressive and anxiety symptoms and deliver cost-effective outcomes (Sarris et al., 2015; Jacka, 2017; Opie et al., 2018; Firth et al., 2019). Mechanistically, diet influences neurotransmitter synthesis, systemic inflammation, and gut-brain signaling, providing biological plausibility.
3. Relevance to payer decision-making.
   These domains are central to employer health plan management, given their cost impact and responsiveness to lifestyle change. By focusing on conditions with both high burden and strong evidence for dietary responsiveness, the analysis provides direct decision-making relevance for payers and benefit consultants (Deana Bell et al., 2024).

**Use of CCSR categories**

Condition definitions were operationalized using Clinical Classifications Software Refined (CCSR) developed by the U.S. Agency for Healthcare Research and Quality (Agency for Healthcare Research and Quality (AHRQ), 2025). CCSR maps ICD-10 codes into clinically meaningful groups, facilitating reproducibility and comparability across studies.

- END009 (Obesity): ICD-10 codes for overweight and obesity, representing the diagnostic group most directly related to metabolic and dietary intervention.
- DIG025 (Functional and other gastrointestinal disorders): Includes IBS, GERD, and related digestive disorders strongly influenced by diet.
- MBD002 (Depressive disorders): Encompasses major depressive disorder, persistent depressive disorder (dysthymia), and other specified or unspecified depressive conditions, all of which contribute significantly to healthcare costs and utilization, and are strongly linked to diet and metabolic health (Lopresti, Hood & Drummond, 2013; Sarris et al., 2015; Firth et al., 2019).
- MBD005 (Anxiety disorders): Includes generalized anxiety disorder, panic disorder, and related conditions where diet has demonstrated measurable clinical and economic impact (Jacka, 2017; Opie et al., 2018).

These categories were selected because they:

- Provide a validated, standardized framework widely used in health services research (Volpp et al., 2023).
- Align precisely with the conditions most responsive to diet-based interventions.
- Map directly to employer-relevant spending categories, ensuring policy relevance.

By applying these CCSR groupings, the analysis remained both clinically specific and methodologically rigorous, while directly targeting the domains most relevant to food-as-medicine strategies and employer health economics.

**Supplementary Note S3. CPT/HCPCS Families and Rationale**

To explain observed changes in medical spend and utilization, the analysis grouped procedure codes that plausibly shift with nutrition-first, metabolic, and behavioral care. We restricted CPT/HCPCS families to cost-relevant services in four study domains—obesity/metabolic, digestive health (IBS/GERD), anxiety, and depressive disorders, plus cross-domain encounter families (ED and E/M). Selections prioritize high-cost or high-volume codes endorsed by clinical guidelines or payer coverage policies, and that are commonly tracked in U.S. medical claims.

**Cross-domain categories (benchmarks for overall use)**

- Emergency department (ED) visits (99281-99285). ED care is a major, potentially preventable cost driver across metabolic, GI, and mental health presentations; tracking ED visits provides a uniform acute-care benchmark (Hsia & Niedzwiecki, 2017).
- Evaluation & Management (E/M) visits (99201-99215, 99221-99233). Office and inpatient E/M reflect routine and hospital-level care intensity and are standard comparators across domains (Centers for Medicare and Medicaid Services, 2025) .

**Obesity**

Rationale. Obesity care spans surgical, nutritional, and behavioral modalities. Claims shifts from invasive or acute care toward guideline-concordant lifestyle and care-management services are expected under food-as-medicine interventions.

- Bariatric procedures (43644, 43645, 43770, 43775, 43845-43847). Bariatric surgery is effective for severe obesity but costly; inclusion captures substitution away from surgical episodes (Arterburn & Courcoulas, 2014; Arterburn et al., 2020).
- Medical nutrition therapy (MNT) (97802-97804; G0270-G0271). Covered by Medicare; RDN-delivered MNT improves cardio-metabolic outcomes and is reimbursed as a distinct service line (Centers for Medicare and Medicaid Services, 2025).
- Intensive behavioral therapy for obesity (G0447; group G0473). National coverage exists for high-intensity behavioral counseling in primary care (Centers for Medicare and Medicaid Services, 2025).
- Diabetes self-management training (G0108-G0109). DSMT is covered and reduces downstream utilization when paired with lifestyle care.
- Metabolic labs (80061 lipid panel; 83036 HbA1c). Recommended in cardiometabolic monitoring.

**Digestive health**

Rationale. Workups for IBS/GERD frequently include repeated endoscopy and physiologic testing; refractory GERD can escalate to surgery. Nutrition-first care may reduce diagnostic churn and procedure intensity.

- Upper endoscopy (EGD) (4323x-4325x; incl. 43239) and colonoscopy (4537x-4539x; incl. 45378). Core diagnostic/therapeutic endoscopy families recommended by ACG guidelines; high contributors to GI spend (Lacy et al., 2021; Katz et al., 2022).
- GERD diagnostics: manometry (91010), ambulatory pH/impedance (91034-91035), hydrogen breath test (91065), esophagram/barium swallow (74220)—all standard tools in reflux and functional GI evaluation (Katz et al., 2022).
- Anti-reflux procedures: fundoplication (43280 laparoscopic; 43327-43328 open), magnetic sphincter augmentation (LINX; 43284), transoral incisionless fundoplication (TIF; 43210), endoscopic radiofrequency (Stretta; 43257), Barrett’s ablation (43229)—included to capture escalation in refractory GERD (Katz et al., 2022)

**Mental health: Anxiety and Depression**

Rationale. Claims commonly reflect stepped care: diagnostic intake → psychotherapy and behavioral services → collaborative care/management → somatic therapies for refractory disease. Diet and metabolic improvement may reduce symptom severity and service intensity, especially urgent/ED use.

- Psychiatric diagnostic evaluation (90791-90792). Standard entry to specialty care.
- Psychotherapy: individual (90832/90834/90837); family (90846-90847); group (90853)—first-line evidence-based treatments for anxiety/depression.
- Psychotherapy add-on with E/M (90833/90836/90838). Captures integrated med-management + therapy encounters (Centers for Medicare and Medicaid Services, 2025).
- Behavioral screening and health behavior services: 96127 (e.g., PHQ-9, GAD-7), 96156, 96158-96159, 96164-96168/96170-96171.
- Collaborative care (CoCM) (99484, 99492-99494). Strong evidence base for improved outcomes and cost-effectiveness in depression/anxiety in primary care (Archer et al., 2012).
- Somatic treatments for depression: ECT (90870-90871) and TMS (90867-90869)—reserved for treatment-resistant cases, high per-episode cost (McClintock et al., 2018).

#### References

Agency for Healthcare Research and Quality (AHRQ). 2025.Clinical Classifications Software Refined (CCSR) for ICD-10-CM Diagnoses. *Available at* *https://hcup-us.ahrq.gov/toolssoftware/ccsr/dxccsr.jsp* (accessed September 30, 2025).

American Diabetes Association Professional Practice Committee, ElSayed NA, McCoy RG, Aleppo G, Balapattabi K, Beverly EA, Briggs Early K, Bruemmer D, Ebekozien O, Echouffo-Tcheugui JB, Ekhlaspour L, Garg R, Khunti K, Lal R, Lingvay I, Matfin G, Pandya N, Pekas EJ, Pilla SJ, Polsky S, Segal AR, Seley JJ, Stanton RC, Bannuru RR. 2025a. 1. Improving Care and Promoting Health in Populations: Standards of Care in Diabetes—2025. *Diabetes Care* 48:S14–S26. DOI: 10.2337/dc25-S001.

American Diabetes Association Professional Practice Committee, ElSayed NA, McCoy RG, Aleppo G, Balapattabi K, Beverly EA, Briggs Early K, Bruemmer D, Echouffo-Tcheugui JB, Ekhlaspour L, Garg R, Khunti K, Kushner RF, Lal R, Lingvay I, Matfin G, Pandya N, Pekas EJ, Pilla SJ, Polsky S, Segal AR, Seley JJ, Stanton RC, Bannuru RR. 2025b. 8. Obesity and Weight Management for the Prevention and Treatment of Type 2 Diabetes: Standards of Care in Diabetes–2025. *Diabetes Care* 48:S167–S180. DOI: 10.2337/dc25-S008.

Archer J, Bower P, Gilbody S, Lovell K, Richards D, Gask L, Dickens C, Coventry P. 2012. Collaborative care for depression and anxiety problems. *Cochrane Database of Systematic Reviews* 2012. DOI: 10.1002/14651858.CD006525.pub2.

Arterburn DE, Courcoulas AP. 2014. Bariatric surgery for obesity and metabolic conditions in adults. *BMJ* 349:g3961–g3961. DOI: 10.1136/bmj.g3961.

Arterburn DE, Telem DA, Kushner RF, Courcoulas AP. 2020. Benefits and Risks of Bariatric Surgery in Adults: A Review. *JAMA* 324:879. DOI: 10.1001/jama.2020.12567.

Berkowitz SA, Terranova J, Randall L, Cranston K, Waters DB, Hsu J. 2019. Association Between Receipt of a Medically Tailored Meal Program and Health Care Use. *JAMA Internal Medicine* 179:786. DOI: 10.1001/jamainternmed.2019.0198.

Centers for Medicare and Medicaid Services. 2025.CPT/HCPCS Codes. *Available at* *https://www.cms.gov/medicare/regulations-guidance/physician-self-referral/list-cpt-hcpcs-codes* (accessed September 30, 2025).

Chatterton ML, Mihalopoulos C, O’Neil A, Itsiopoulos C, Opie R, Castle D, Dash S, Brazionis L, Berk M, Jacka F. 2018. Economic evaluation of a dietary intervention for adults with major depression (the “SMILES” trial). *BMC Public Health* 18:599. DOI: 10.1186/s12889-018-5504-8.

Deana Bell, Clarkson Jason, Mike Gaal, Dave Liner, Annie Man, Andrew Naugle. 2024.2024 Milliman Medical Index. *Available at* *https://www.milliman.com/en/insight/2024-milliman-medical-index* (accessed September 30, 2025).

Deng S, Hager K, Wang L, Cudhea FP, Wong JB, Kim DD, Mozaffarian D. 2025. Estimated Impact Of Medically Tailored Meals On Health Care Use And Expenditures In 50 US States: Article examines the impact of medically tailored meals on health care use and expenditures in 50 US states. *Health Affairs* 44:433–442. DOI: 10.1377/hlthaff.2024.01307.

Firth J, Marx W, Dash S, Carney R, Teasdale SB, Solmi M, Stubbs B, Schuch FB, Carvalho AF, Jacka F, Sarris J. 2019. The Effects of Dietary Improvement on Symptoms of Depression and Anxiety: A Meta-Analysis of Randomized Controlled Trials. *Psychosomatic Medicine* 81:265–280. DOI: 10.1097/PSY.0000000000000673.

Hall ME, Cohen JB, Ard JD, Egan BM, Hall JE, Lavie CJ, Ma J, Ndumele CE, Schauer PR, Shimbo D, on behalf of the American Heart Association Council on Hypertension; Council on Arteriosclerosis, Thrombosis and Vascular Biology; Council on Lifestyle and Cardiometabolic Health; and Stroke Council. 2021. Weight-Loss Strategies for Prevention and Treatment of Hypertension: A Scientific Statement From the American Heart Association. *Hypertension* 78. DOI: 10.1161/HYP.0000000000000202.

Halmos EP, Power VA, Shepherd SJ, Gibson PR, Muir JG. 2014. A Diet Low in FODMAPs Reduces Symptoms of Irritable Bowel Syndrome. *Gastroenterology* 146:67-75.e5. DOI: 10.1053/j.gastro.2013.09.046.

Hsia RY, Niedzwiecki M. 2017. Avoidable emergency department visits: a starting point. *International Journal for Quality in Health Care* 29:642–645. DOI: 10.1093/intqhc/mzx081.

Jacka FN. 2017. Nutritional Psychiatry: Where to Next? *EBioMedicine* 17:24–29. DOI: 10.1016/j.ebiom.2017.02.020.

Katz PO, Dunbar KB, Schnoll-Sussman FH, Greer KB, Yadlapati R, Spechler SJ. 2022. ACG Clinical Guideline for the Diagnosis and Management of Gastroesophageal Reflux Disease. *American Journal of Gastroenterology* 117:27–56. DOI: 10.14309/ajg.0000000000001538.

Kwon OY, Lee MK, Lee HW, Kim H, Lee JS, Jang Y. 2024. Mobile App–Based Lifestyle Coaching Intervention for Patients With Nonalcoholic Fatty Liver Disease: Randomized Controlled Trial. *Journal of Medical Internet Research* 26:e49839. DOI: 10.2196/49839.

Lacy BE, Pimentel M, Brenner DM, Chey WD, Keefer LA, Long MD, Moshiree B. 2021. ACG Clinical Guideline: Management of Irritable Bowel Syndrome. *American Journal of Gastroenterology* 116:17–44. DOI: 10.14309/ajg.0000000000001036.

Liu L, Wang H, Chen X, Zhang Y, Zhang H, Xie P. 2023. Gut microbiota and its metabolites in depression: from pathogenesis to treatment. *eBioMedicine* 90:104527. DOI: 10.1016/j.ebiom.2023.104527.

Lopresti AL, Hood SD, Drummond PD. 2013. A review of lifestyle factors that contribute to important pathways associated with major depression: Diet, sleep and exercise. *Journal of Affective Disorders* 148:12–27. DOI: 10.1016/j.jad.2013.01.014.

McClintock SM, Reti IM, Carpenter LL, McDonald WM, Dubin M, Taylor SF, Cook IA, O’Reardon J, Husain MM, Wall C, Krystal AD, Sampson SM, Morales O, Nelson BG, Latoussakis V, George MS, Lisanby SH, on behalf of both the National Network of Depression Centers rTMS Task Group and the American Psychiatric Association Council on Research Task Force on Novel Biomarkers and Treatments. 2018. Consensus Recommendations for the Clinical Application of Repetitive Transcranial Magnetic Stimulation (rTMS) in the Treatment of Depression: (Consensus Statement). *The Journal of Clinical Psychiatry* 79:35–48. DOI: 10.4088/JCP.16cs10905.

Opie RS, O’Neil A, Jacka FN, Pizzinga J, Itsiopoulos C. 2018. A modified Mediterranean dietary intervention for adults with major depression: Dietary protocol and feasibility data from the SMILES trial. *Nutritional Neuroscience* 21:487–501. DOI: 10.1080/1028415X.2017.1312841.

Ridberg R, Sharib JR, Garfield K, Hanson E, Mozaffarian D. 2025. ‘Food Is Medicine’ In The US: A National Survey Of Public Perceptions Of Care, Practices, And Policies: Article examines the results of a national survey of US public awareness and perceptions of Food Is Medicine interventions. *Health Affairs* 44:398–405. DOI: 10.1377/hlthaff.2024.00585.

Sandhu DS, Fass R. 2018. Current Trends in the Management of Gastroesophageal Reflux Disease. *Gut and Liver* 12:7–16. DOI: 10.5009/gnl16615.

Sarris J, Logan AC, Akbaraly TN, Amminger GP, Balanzá-Martínez V, Freeman MP, Hibbeln J, Matsuoka Y, Mischoulon D, Mizoue T, Nanri A, Nishi D, Ramsey D, Rucklidge JJ, Sanchez-Villegas A, Scholey A, Su K-P, Jacka FN. 2015. Nutritional medicine as mainstream in psychiatry. *The Lancet Psychiatry* 2:271–274. DOI: 10.1016/S2215-0366(14)00051-0.

Shah ED, Salwen-Deremer JK, Gibson PR, Muir JG, Eswaran S, Chey WD. 2021. Pharmacologic, Dietary, and Psychological Treatments for Irritable Bowel Syndrome With Constipation: Cost Utility Analysis. *MDM Policy & Practice* 6:2381468320978417. DOI: 10.1177/2381468320978417.

Shah ED, Salwen-Deremer JK, Gibson PR, Muir JG, Eswaran S, Chey WD. 2022. Comparing Costs and Outcomes of Treatments for Irritable Bowel Syndrome With Diarrhea: Cost-Benefit Analysis. *Clinical Gastroenterology and Hepatology* 20:136-144.e31. DOI: 10.1016/j.cgh.2020.09.043.

Volpp KG, Berkowitz SA, Sharma SV, Anderson CAM, Brewer LC, Elkind MSV, Gardner CD, Gervis JE, Harrington RA, Herrero M, Lichtenstein AH, McClellan M, Muse J, Roberto CA, Zachariah JPV, on behalf of the American Heart Association. 2023. Food Is Medicine: A Presidential Advisory From the American Heart Association. *Circulation* 148:1417–1439. DOI: 10.1161/CIR.0000000000001182.

Zalvan CH, Hu S, Greenberg B, Geliebter J. 2017. A Comparison of Alkaline Water and Mediterranean Diet vs Proton Pump Inhibition for Treatment of Laryngopharyngeal Reflux. *JAMA Otolaryngology–Head & Neck Surgery* 143:1023. DOI: 10.1001/jamaoto.2017.1454.
