## Supplementary material for "Economic Impact of a Precision Nutrition Digital Therapeutic on Employer Health Costs: A Multi-Employer and Multi-Year Claims Analysis": Supp Table S1

**Supplementary Table S1.** Association of program enrollment with healthcare spending by CPT/HCPCS family, stratified by disease domain.

Estimates are presented on the log scale with 95% confidence intervals (CI), alongside the corresponding percentage change and absolute per-member-per-month (PMPM) change in costs. Negative values represent reductions in spending. The table reports results only for CPT families with sufficient sample size and relevance in each disease category. *p-values* are based on regression models with treatment-control comparisons in the 12-month pre/post window. "Enrolled (N)" refers to the number of treated members with at least one claim in the category, "Never-Enrolled (N)" refers to matched controls, and "Employers (N)" reflects the number of participating employer groups contributing data for that cell.

| **Cost Category** | **CPT/HCPCS**  **Codes Family** | **Estimate (95% CI, log scale)** | **% Change (95% CI)** | **PMPM Change (95% CI)** | **p-value** | **Enrolled (N)** | **Never-Enrolled (N)** | **Employers (N)** |
| --- | --- | --- | --- | --- | --- | --- | --- | --- |
| Obesity | | | | | | | | |
|  | Office EM | -0.127 (-0.456, 0.202) | -12 (-37, 22) | -17 (-53, 32) | 0.449 | 13 | 568 | 3 |
| Digestive Health | | | | | | | | |
|  | Medical nutrition therapy | -2.653 (-3.786, -1.519) | -93 (-98, -78) | -283 (-297, -238) | 5.41E-06 | 8 | 158 | 1 |
| Mental Health | | | | | | | | |
|  | Psychotherapy individual | -0.054 (-0.266, 0.158) | -5 (-23, 17) | -15 (-68, 50) | 0.617 | 29 | 708 | 3 |
|  | Psychotherapy addon with EM | -0.206 (-0.358, -0.055) | -19 (-30, -5) | -21 (-34, -6) | 0.008 | 8 | 149 | 1 |
| Anxiety | | | | | | | | |
|  | Psychotherapy individual | 0.293 (-0.128, 0.714) | 34 (-12, 104) | 91 (-32, 279) | 0.173 | 9 | 216 | 2 |
|  | Psychotherapy addon with EM | -0.172 (-0.269, -0.076) | -16 (-24, -7) | -18 (-27, -8) | 5.18E-04 | 6 | 107 | 1 |
| Depression | | | | | | | | |
|  | Psychotherapy individual | -0.305 (-0.965, 0.354) | -26 (-62, 42) | -77 (-182, 125) | 0.364 | 6 | 129 | 2 |
|  | Psychotherapy addon with EM | -0.226 (-0.423, -0.029) | -20 (-34, -3) | -23 (-39, -3) | 0.025 | 6 | 77 | 1 |
