## Supplementary material for "Economic Impact of a Precision Nutrition Digital Therapeutic on Employer Health Costs: A Multi-Employer and Multi-Year Claims Analysis": Supp Table S2

**Supplementary Table SXX**. Baseline characteristics and spending by employer and group (treated vs. control).

This table reports employer-level baseline descriptive statistics for treated (members who enrolled in the digital health intervention) and never-enrolled control members. For each employer and group the table reports: number of unique members (N Members), mean age (SD), percent female (SD), mean number of pre-enrolment months observed (SD), and mean (SD) per-member dollar spending in U.S. dollars for: Total Medical spending, Total Diet-responsive (controllable) spending, Obesity, Digestive Disorders, and Mental Health (Anxiety + Depression). All spending values are expressed as mean (SD) for the baseline window described below and are conditional on months with employer-paid claims (see Methods).

Standardized mean differences (SMD): For each employer we report the standardized mean difference (SMD) comparing treated and control members on each variable. For continuous variables SMD was computed as the difference in group means divided by the pooled standard deviation. For the binary variable (% female) SMD was computed as $(p_{t}-p_{c})/\sqrt{(p_{t}(1-p_{t})+p_{c}(1-p_{c}))/2}$. Absolute SMD values are reported; as a guide, $\mid\text{SMD}\mid<0.10$indicates negligible imbalance, $0.10\leq\mid\text{SMD}\mid<0.20$small imbalance, $0.20\leq\mid\text{SMD}\mid<0.50$moderate imbalance, and $\mid\text{SMD}\mid\geq0.50$large imbalance.

Baseline window and PMPM definition: For treated members the baseline period comprises months with rel_time < 0 (the months prior to enrollment); for controls, baseline observations were restricted to never-enrolled members observed in the same partner × calendar months that appear in treated members’ pre-periods so that the calendar-month composition is comparable. The spending metrics shown are per-member per-month means conditional on months with positive employer-paid spending (i.e., months with paid claims only), as described in Methods.

**Interpretive notes and caveats.** Large absolute SMDs indicate important baseline differences between treated and control members within that employer and should be interpreted with caution. Several employer strata have very small treated sample sizes (for example, treated N ≤ 5), in which case the SMD and mean estimates are unstable. Importantly, our event-study diagnostics (Supplementary Figure S1) indicate no systematic pre-treatment trends for the pooled sample, and the DiD2S first stage removes time-invariant individual-level differences. Abbreviations. SD = standard deviation; SMD = standardized mean difference; PMPM = per-member per-month. Values shown as *mean (SD)*. See Methods 2.2–2.5 for full variable definitions.

|  |  |  |  |  |  | **Baseline Spent by Category (USD$)** | | | | |
| --- | --- | --- | --- | --- | --- | --- | --- | --- | --- | --- |
| **Employer identifier** | **Treatment Group** | **N Members** | **Age** | **% Female** | **N Months pre** | **Total Medical** | **Total Diet-responsive** | **Obesity** | **Digestive Disorders** | **Mental Health (Anxiety + Depression)** |
| **36** | **Control** | 1192 | 50.3 (12.8) | 57.3 (49.2) | 5.4 (6.4) | 757 (2658) | 739 (2690) | 1385 (5736) | 0(0) | 199 (511) |
|  | **Treated** | 43 | 51.4 (9.2) | 58.7 (49.2) | 4 (3.5) | 1433 (5984) | 1438 (6189) | 440 (552) | 0(0) | 159 (101) |
|  | **SMD** |  | -0.09 | -0.03 | 0.23 | -0.24 | -0.24 | 0.17 |  | 0.08 |
| **138** | **Control** | 642 | 45.1 (13.7) | 52.2 (50) | 3.2 (2.7) | 1796 (6042) | 1448 (8938) | 0 (0) | 0(0) | 487 (3593) |
|  | **Treated** | 8 | 44.1 (12.8) | 62.5 (51.8) | 2 (2.1) | 2950 (4485) | 515 (1034) | 0 (0) | 0(0) | 145 (34) |
|  | **SMD** |  | 0.07 | -0.21 | 0.44 | -0.19 | 0.10 |  |  | 0.10 |
| **168** | **Control** | 2895 | 44.5 (13.4) | 53 (49.9) | 2.9 (2.4) | 1754 (6463) | 862 (3281) | 404 (1288) | 868(3564) | 385 (1072) |
|  | **Treated** | 147 | 43.8 (9.9) | 58.8 (49.4) | 1.7 (1.4) | 1805 (7506) | 920 (1924) | 190 (107) | 493(406) | 564 (1315) |
|  | **SMD** |  | 0.05 | -0.12 | 0.50 | -0.01 | -0.02 | 0.17 | 0.11 | -0.17 |
| **174** | **Control** | 603 | 52.2 (12.5) | 41.7 (45.4) | 1.7 (1.1) | 569 (1310) | 558 (1263) | 0 (0) | 0(0) | 0 (0) |
|  | **Treated** | 3 | 64 (22.1) | 66.7 (57.7) | 3.3 (4) | 228 (256) | 228 (256) | 0 (0) | 0(0) | 0 (0) |
|  | **SMD** |  | -0.94 | -0.55 | -1.46 | 0.26 | 0.26 |  |  |  |
| **186** | **Control** | 1647 | 46 (13.8) | 57.2 (49.4) | 2.9 (2.4) | 1863 (5642) | 1067 (3704) | 1260 (6846) | 2336(15397) | 543 (1754) |
|  | **Treated** | 22 | 49.5 (9.4) | 80 (40.8) | 1.5 (1.1) | 858 (2389) | 913 (2592) | 103 (75) | 356(317) | 182 (153) |
|  | SMD |  | -0.25 | -0.46 | 0.61 | 0.18 | 0.04 | 0.17 | 0.13 | 0.21 |
| 192.0 | Control | 1236 | 50.5 (14) | 61.9 (45.2) | 3.3 (2.5) | 1600 (7002) | 980 (4562) | 1653 (11409) | 0(0) | 375 (2087) |
|  | Treated | 31 | 49.4 (10.3) | 87.6 (33.1) | 3.6 (2.4) | 1037 (2628) | 421 (904) | 263 (32) | 0(0) | 156 (131) |
|  | SMD |  | 0.08 | -0.57 | -0.16 | 0.08 | 0.12 | 0.12 |  | 0.11 |
| 210.0 | Control | 150 | 50.1 (15) | 54 (50) | 1.2 (0.4) | 6905 (15936) | 3348 (7540) | 0 (0) | 0(0) | 0 (0) |
|  | Treated | 4 | 53.5 (11.6) | 50 (57.7) | 1.3 (0.5) | 11548 (12106) | 10544 (10835) | 0 (0) | 0(0) | 0 (0) |
|  | SMD |  | -0.23 | 0.08 | -0.02 | -0.29 | -0.94 |  |  |  |
