## Supplementary material for "Economic Impact of a Precision Nutrition Digital Therapeutic on Employer Health Costs: A Multi-Employer and Multi-Year Claims Analysis": Supp Table S4

**Supplementary Table S2**. Association of program enrollment with healthcare utilization by CPT/HCPCS family, stratified by disease domain.

Estimates are reported as log-transformed changes in utilization with 95% confidence intervals (CI), converted to relative percentage change and absolute change in mean visits or procedures per member per month (PMPM). Negative values indicate reductions in utilization. Results are presented only for CPT families with adequate sample size and relevance within each disease category. p-values are derived from regression models comparing treated versus never-enrolled controls across the 12-month pre/post observation window. "Enrolled (N)" represents the number of treated members contributing at least one claim in the category, "Never-Enrolled (N)" denotes matched controls, and "Employers (N)" indicates the number of participating employer groups represented in that category.

| **Cost Category** | **CPT/HCPCS**  **Codes Family** | **Estimate (95% CI, log scale)** | **% Change (95% CI)** | **PMPM Change (95% CI)** | **p-value** | **Enrolled (N)** | **Never-Enrolled (N)** | **Employers (N)** |
| --- | --- | --- | --- | --- | --- | --- | --- | --- |
| Obesity | | | | | | | | |
|  | Office EM | -0.241 (-0.424, -0.057) | -21 (-35, -6) | -0.25 (-0.4, -0.06) | 0.010 | 13 | 568 | 3 |
| Digestive Health | | | | | | | | |
|  | Medical nutrition therapy | -0.035 (-0.085, 0.014) | -3 (-8, 1) | -0.04 (-0.08, 0.01) | 0.162 | 8 | 158 | 1 |
| Mental Health | | | | | | | | |
|  | Psychotherapy individual | -0.065 (-0.279, 0.149) | -6 (-24, 16) | -0.15 (-0.57, 0.38) | 0.549 | 29 | 708 | 3 |
|  | Psychotherapy addon with EM | -0.044 (-0.127, 0.04) | -4 (-12, 4) | -0.05 (-0.14, 0.05) | 0.302 | 8 | 149 | 1 |
| Anxiety | | | | | | | | |
|  | Psychotherapy individual | 0.163 (-0.203, 0.528) | 18 (-18, 70) | 0.4 (-0.41, 1.57) | 0.382 | 9 | 216 | 2 |
|  | Psychotherapy addon with EM | 0.025 (-0.099, 0.15) | 3 (-9, 16) | 0.03 (-0.11, 0.19) | 0.690 | 6 | 107 | 1 |
| Depression | | | | | | | | |
|  | Psychotherapy individual | -0.421 (-1.229, 0.386) | -34 (-71, 47) | -0.8 (-1.64, 1.09) | 0.306 | 6 | 129 | 2 |
|  | Psychotherapy addon with EM | -0.021 (-0.142, 0.1) | -2 (-13, 10) | -0.02 (-0.16, 0.12) | 0.731 | 6 | 77 | 1 |
